# Effect of a novel food rich in miraculin on the intestinal microbiome of malnourished patients with cancer and dysgeusia

**DOI:** 10.1101/2024.08.20.24312287

**Authors:** Julio Plaza-Diaz, Marco Brandimonte-Hernández, Bricia López-Plaza, Francisco Javier Ruiz-Ojeda, Ana Isabel Álvarez-Mercado, Lucía Arcos-Castellanos, Jaime Feliú-Batlle, Thomas Hummel, Samara Palma Milla, Angel Gil

**Author notes:** These authors contributed equally to this work (Samara Palma Milla, Angel Gil). Correspondence (A.G.); (A.G.).

## Abstract

Dysgeusia contributes to nutritional derangement and worsens the quality of life of patients with cancer. Despite the different strategies, there is no effective treatment for patients suffering from taste disorders provided by the pharmaceutical industry. We developed a novel strategy for reducing side effects in cancer patients by providing a novel food supplement with the taste-modifying glycoprotein miraculin, which is approved by the European Union, as an adjuvant to medical-nutritional therapy. A pilot randomized, parallel, triple-blind, and placebo-controlled intervention clinical trial was carried out in which 31 malnourished patients with cancer and dysgeusia receiving antineoplastic treatment, and were randomized into three arms: standard dose of DMB (150 mg DMB/tablet), high dose of DMB (300 mg DMB/tablet) or placebo (300 mg freeze-dried strawberry) for three months. Patients consumed a DMB or placebo tablet before each main meal (breakfast, lunch and dinner). Using stool samples from patients with cancer, we analyzed the intestinal microbiome via nanopore methodology. We detected differences in the relative abundances of genera *Phocaeicola* and *Escherichia* depending on the treatment. Nevertheless, only the *Solibaculum* genus was more abundant in the standard-dose DMB group after 3 months. At the species level, *Bacteroides* sp. PHL 2737 presented a relatively low abundance in both DMB groups, and *Vescimonas coprocola* presented a relatively high abundance in both treatment groups after 3 months. Furthermore, a standard dose of DMB was positively associated with TNF-α levels and *Lachnoclostridium* and *Mediterraneibacter* abundances, whereas a high dose of DMB was negatively associated with TNF-α levels and the relative abundance of *Phocaeicola*. After a high dose of DMB, erythrocyte polyunsaturated fatty acids were positively correlated with *Lachnoclostridium* and *Roseburia*, and there was a positive association between *Phocaeicola* and the acetic acid concentration of feces. The intake of DMB together with nutritional treatment and individualized dietary advice results in positive changes in the intestinal microbiome of patients with cancer and dysgeusia There was a negative association between the relative abundance of *Phocaeicola* and taste perception in the DMB high dose group. Changes observed in the intestinal microbiota might contribute to maintaining an appropriate immune response of cancer patients. Since the present pilot study involved only a few participants, further research is needed to draw robust conclusions.

## 1. Introduction

Cancer is characterized by uncontrolled cell proliferation [1]. The disease affects people in many ways, including psychologically, physically, economically, and socially [2]. Many patients with cancer may benefit from systemic therapy, chemotherapy, and radiotherapy; however, these treatments are also associated with a high risk of serious complications [3].

Malnutrition is estimated to be responsible for the death of 10-20% of patients with cancer [4]. However, nutritional support is received by only 30%-60% of cancer patients who are at risk of malnutrition [4].

Despite possible adverse consequences, taste changes experienced by patients with cancer are not usually diagnosed and treated early because clinicians do not consider them life-threatening [5-7]. It is estimated that 45 to 80% of patients with chemotherapy-induced taste changes will experience these changes [8-10]. Dysgeusia is the umbrella term for qualitative and quantitative taste dysfunction, and includes taste distortions with bitter, metallic, salty, or unpleasant tastes [11-13]. The consequences of taste alterations are the deterioration of nutritional status, a reduction in quality of life, weight loss, and ultimately, health [14-17]. Zinc, amifostine, selenium, lactoferrin, and cannabinoids are currently used to treat taste disorders; however, their effectiveness is limited [18-20].

The gut microbiota plays a crucial role in maintaining health, influencing not only the gastrointestinal tract, but also distant organs such as the brain, liver, and pancreas [21,22]. The composition of the gut microbiota is diverse: it is composed of more than 200 bacterial species [23,24] (including phylotypes such as *Bacillota, Bacteroidota, Actinomyces, Fusobacterium, Pseudomonadota, and Verrucomicrobiota*) [25], fungi (*Candida albicans*), viruses and protists [26]. Microorganisms that belong to a separate kingdom of living organisms, *Archaebacteria*, are also an important part of the intestinal microbiota [27]. An alteration in the equilibrium of the gut microbiota can result in the development of a dysbiotic state, with subsequent implications for both local and systemic health outcomes [28]. Thus, dysbiosis contributes to a variety of pathologies, including obesity [29], diabetes [30], neurodegenerative diseases [31], and cancer [32]. Approximately 20% of all cancers are strongly associated with specific viral or microbial infections [33]. Furthermore, bacteria have been identified as key factors in the progression of several types of cancer, including oral squamous cell carcinomas, colorectal cancer, and pancreatic ductal adenocarcinoma [34-36].

The complexity of the gut microbiome, as well as its richness and abundance, predicts the metabolic health of the host [37]. Several factors contribute to the composition of the gut microbiome, including diet and dietary habits. Unsurprisingly, the gut microbiome has been associated with several cancer determinants, such as taste perception, which influences appetite regulation and energy metabolism [37]. Furthermore, there is evidence that the gut microbiota can affect the response to systemic cancer therapy [38].

Miraculin is a glycoprotein obtained from the *Synsepalum dulcificum* berries that converts a sour taste into a sweet taste, which is why the fruit is also called “miracle berry” [39]. The taste-modifying effect of miraculin occurs under acidic conditions and lasts for approximately 30 minutes after consumption. Two small non-randomized studies using non-objective tools tried to evaluate the effect of miracle fruit on taste disorders in patients with cancer who are receiving active chemotherapy treatment describing promising results [15,40]. Dried miracle berries (DMB) was approved as a novel food by the European Commission in December 2021. In addition to its taste-modifying properties, DMB also contains bioactive ingredients, such as fiber and phenolic compounds [41,42].

In a pilot randomized, parallel, triple-blind, and placebo-controlled clinical trial (the CLINMIR study), our research group provided clinical evidence on the efficacy of DMB in improving taste alterations in cancer patients. As a result of this study, we observed improvements in electrochemical food perception, energy and nutrient intake, nutritional status, and quality of life for malnourished patients with cancer receiving antineoplastic treatment [43]. Moreover, we showed that regular consumption of DMB consumption and nutritional interventions changed the oral microbiome in patients with cancer and dysgeusia, which may contribute to maintaining an appropriate immune response without altering taste perception [44]. The purpose of the present study was to assess the intestinal microbiome of malnourished patients with cancer and dysgeusia after DMB consumption as a medical-nutritional adjuvant treatment.

## 2. Results

During the period of November 2022 to May 2023, 62 patients were assessed for eligibility. Among 31 patients with cancer who met the inclusion criteria, three intervention groups were randomly assigned according to the type of cancer. In the course of the study, ten participants withdrew from the study. Several of these dropouts were caused by taste distortions caused by acidic foods that were not sweet (n = 6) and the complexity of the intervention prescription (n = 2). During the course of the study, two placebo patients died. A total of 21 patients with cancer completed the clinical trial. All variables were analyzed with an intention-to-treat approach. The sample consisted of 58.1% women and 41.9% men, with an average age of 60.0 ± 10.9 years. Participants who were actively treated were assessed by electrogustometry; results of taste perception for the population have been published elsewhere [45].

### 2.1. Phylum level

At baseline, *Bacillota* and *Bacteroidota* accounted for more than 80% of the relative abundance of the intestinal microbiota. Based on the comparison between baseline and three months, no major differences were found among the groups. According to the treatment, only *Pseudomonadota* was significantly different among the three study groups. Both the alpha diversity indices (Shannon, Simpson and Chao1) and the studied phyla did not show any effect of treatment x time (Table 1).

**Table 1.** Relative abundances of intestinal bacteria at the phylum level in malnourished patients with cancer and dysgeusia who received standard-dose DMB (150 mg), high-dose DMB (300 mg) or placebo for 3 months.

| Phylum | DMB 150 mg |  | DMB 300 mg |  | Placebo |  | p-value |  |  |
| --- | --- | --- | --- | --- | --- | --- | --- | --- | --- |
|  | Baseline | 3 months | Baseline | 3 months | Baseline | 3 months | Treatment<br>(T) | Time<br>(t) | T x t |
| *Actinobacteriota* | 0.3 | 0.4 | 0.2 | 0.2 | 0.3 | 0.1 | 0.717 | 0.357 | 0.468 |
|  | (0.1 - 0.7) | (0.1 - 0.4) | (0.03 - 0.7) | (0.07 - 0.5) | (0.01 - 0.8) | (0.03 - 0.6) |  |  |  |
| *Bacillota* | 78.1 | 88.6 | 75.7 | 74.7 | 69.1 | 73.7 | 0.062 | 0.224 | 0.598 |
|  | (70.6 - 94.1) | (76 - 95.1) | (49.3 - 86) | (67.7 - 80.6) | (13.1 - 88.1) | (50.3 - 91.1) |  |  |  |
| *Bacteroidota* | 14.6 | 8.2 | 8.9 | 16 | 10.2 | 6.7 | 0.444 | 0.674 | 0.888 |
|  | (1.4 - 24.5) | (1.9 - 20.7) | (3.1 - 41.4) | (3 - 26.4) | (0.008 - 18.1) | (0.1 - 32.4) |  |  |  |
| *Pseudomonadota* | 3.8 | 2 | 7.2 | 8.1 | 16.8 | 15.9 | 0.043* | 0.253 | 0.366 |
|  | (2.2 - 7.9) | (1.2 - 12) | (4.6 - 24) | (3.4 - 21.5) | (2.7 - 86.8) | (4.2 - 31.4) |  |  |  |
| *Tenericutes* | 0.2 | 0.06 | 0.09 | 0.03 | 0.1 | 0.06 | 0.542 | 0.092 | 0.703 |
|  | (0.01 - 0.3) | (0.01 - 0.2) | (0.01 - 0.2) | (0.008 - 0.2) | (0.006 - 0.4) | (0.01 - 0.2) |  |  |  |
| *Synergistetes* | 0.1 | 0.08 | 0.08 | 0.2 | 0.2 | 0.1 | 0.282 | 0.397 | 0.876 |
|  | (0.008 - 0.3) | (0.01 - 0.2) | (0.01 - 1.3) | (0.02 - 0.5) | (0.05 - 0.2) | (0.01 - 0.2) |  |  |  |
| *Verrucomicrobiota* | 0.1 | 0.1 | 0.03 | 0.05 | 0.08 | 0.04 | 0.616 | 0.466 | 0.388 |
|  | (0.006 - 0.9) | (0.01 - 1.5) | (0.007 - 1.3) | (0.01 - 0.2) | (0.07 - 1.2) | (0.01 - 2.4) |  |  |  |
| Shannon | 3.3 | 3.3 | 3.3 | 3.3 | 3.3 | 3.0 | 0.150 | 0.879 | 0.745 |
|  | (2.5 - 3.6) | (3.1 - 3.8) | (2.3 - 3.7) | (2.6 - 3.4) | (1.5 - 3.3) | (2.1 - 3.8) |  |  |  |
| Simpson | 0.9 | 0.9 | 0.9 | 0.9 | 0.9 | 0.9 | 0.135 | 0.825 | 0.579 |
|  | (0.8 - 0.9) | (0.9 - 1.0) | (0.8 - 0.9) | (0.9 - 0.9) | (0.5 - 0.9) | (0.8 - 1.0) |  |  |  |
| Chao1 | 435.2 | 388.7 | 404.7 | 422.6 | 394.7 | 415.6 | 0.367 | 0.139 | 0.202 |
|  | (308.3 - 548.2) | (315.0 - 540.0) | (255.1 - 684.8) | (261.0 - 491.2) | (264.3 - 514.2) | (272.1 - 547.0) |  |  |  |

The values are presented as medians and ranges. Based on the median test across time points within groups, \*indicates significant differences (p<0.05).

### 2.2. Genus level

The most common genus in all the studied groups was *Faecalibacterium* (approximately 11 to almost 20 % relative abundance). There were differences between the genera *Phocaeicola* and *Escherichia* depending on the treatment. For *Solibaculum*, we observed significant differences in the interaction effect of treatment x time. The standard dose of DMB produced a significant increase in the relative abundance of *Solibaculum*, whereas the placebo resulted in a significant decrease in the relative abundance of this genus (Table 2).

**Table 2.** Relative abundances of intestinal bacteria at the genus level in malnourished patients with cancer and dysgeusia who received standard-dose DMB (150 mg), high-dose DMB (300 mg) or placebo for 3 months.

| Genus | DMB 150 mg |  | DMB 300 mg |  | Placebo |  | p-value |  |  |
| --- | --- | --- | --- | --- | --- | --- | --- | --- | --- |
|  | First visit | 3 months | First visit | 3 months | First visit | 3 months | Treatment<br>(T) | Time<br>(t) | T × t |
| *Faecalibacterium* | 17.5<br>(5.7 - 34.1) | 13.4<br>(12 - 24.2) | 19.7<br>(4.6 - 28.8) | 17.2<br>(4.6 - 28.8) | 11.4<br>(0.01 - 35) | 12.2<br>(0.05 - 46.3) | 0.614 | 0.707 | 0.670 |
| *Prevotella* | 0.04<br>(0.01 - 10.8) | 0.4<br>(0.01 - 15.8) | 1.7<br>(0.02 - 30.9) | 5.6<br>(0.1 - 23.8) | 3.9<br>(0.01 - 15.5) | 3.7<br>(0.1 - 27.9) | 0.669 | 0.936 | 0.722 |
| *Blautia* | 4.2<br>(2.4 - 8.4) | 4.9<br>(2.3 - 7.8) | 4.7<br>(0.5 - 10.1) | 3.9<br>(2.3 - 7.5) | 2.9<br>(0.02 - 9) | 2.9<br>(0.02 - 4.4) | 0.382 | 0.236 | 0.828 |
| *Anaerobutyricum* | 4.0<br>(1 - 7.8) | 2.4<br>(1.6 - 5.4) | 2.6<br>(0.2 - 6.5) | 2.3<br>(1.2 - 5.3) | 1.9<br>(0.5 - 2.9) | 3<br>(0.01 - 4.2) | 0.369 | 0.848 | 0.126 |
| *Dysosmobacter* | 3.3<br>(0.8 - 4) | 3.7<br>(0.7 - 6.3) | 1.5<br>(0.5 - 7.1) | 2.9<br>(0.3 - 9.3) | 2.6<br>(1.3 - 3.8) | 2.6<br>(0.02 - 6.2) | 0.841 | 0.299 | 0.859 |
| *Vescimonas* | 3.3<br>(0.2 - 10.6) | 3.3<br>(1.1 - 18.8) | 2.6<br>(0.1 - 8.6) | 1.3<br>(0.03 - 8.4) | 6<br>(0.5 - 17.4) | 2.8<br>(0.01 - 12.6) | 0.450 | 0.977 | 0.130 |
| *Roseburia* | 2.8<br>(0.7 - 21.4) | 1.7<br>(0.9 - 23.4) | 2.4<br>(1.2 - 13.2) | 3.6<br>(0.4 - 11.6) | 1.6<br>(1.4 - 4.9) | 3.2<br>(1.6 - 12.4) | 0.830 | 0.506 | 0.547 |
| *Sulcia* | 2.8<br>(2.8 - 2.8) | 1.1<br>(1.1 - 1.1) | 0<br>(0 - 0) | 0<br>(0 - 0) | 0.3<br>(0.3 - 0.3) | 0<br>(0 - 0) | 1.0 | 1.0 | 1.0 |
| *Bacteroides* | 2.5<br>(0.4 - 9.4) | 1.3<br>(0.7 - 4.8) | 1.9<br>(0.8 - 14) | 2.2<br>(0.6 - 8) | 0.9<br>(0.3 - 15.5) | 0.7<br>(0.02 - 7.7) | 0.915 | 0.131 | 0.957 |
| *Lachnospira* | 2.1<br>(1.3 - 4) | 2.5<br>(0.5 - 3.4) | 2.5<br>(0.4 - 3.4) | 1.3<br>(0.3 - 3.8) | 1.1<br>(0.6 - 10.8) | 1.8<br>(0.6 - 6.4) | 0.576 | 0.459 | 0.874 |
| *Clostridium* | 1.8<br>(1.2 - 2.4) | 1.9<br>(0.9 - 4.3) | 2.1<br>(0.2 - 4.1) | 1.3<br>(0.9 - 16.2) | 1.2<br>(0.9 - 3.1) | 1.4<br>(0.7 - 40.1) | 0.614 | 0.182 | 0.513 |
| *Coprococcus* | 1.5<br>(1 - 3.6) | 2.2<br>(0.8 - 2.8) | 0.9<br>(0.06 - 5) | 1.1<br>(0.2 - 2.2) | 1<br>(0.2 - 2.7) | 1.9<br>(0.5 - 2.9) | 0.558 | 0.630 | 0.590 |
| *Blattabacterium* | 1.5<br>(1.5 - 1.5) | 0.5<br>(0.5 - 0.5) | 0<br>(0 - 0) | 0<br>(0 - 0) | 0.2<br>(0.2 - 0.2) | 0<br>(0 - 0) | 1.0 | 1.0 | 1.0 |
| *Phascolarctobacterium* | 1.4<br>(0.2 - 3) | 1.9<br>(1.4 - 10.8) | 2<br>(1.1 - 2.6) | 1.6<br>(0.08 - 6.7) | 0.03<br>(0.007 - 8.4) | 0.01<br>(0.006 - 9.2) | 0.966 | 0.206 | 0.586 |
| *Mediterraneibacter* | 1.3<br>(0.5 - 3.3) | 0.8<br>(0.4 - 2.5) | 1.6<br>(0.3 - 9) | 1.5<br>(0.4 - 4.5) | 0.7<br>(0.5 - 1.5) | 0.8<br>(0.03 - 4.8) | 0.379 | 0.514 | 0.350 |
| *Dorea* | 1.2<br>(0.9 - 2.9) | 1.3<br>(0.8 - 2.8) | 2.3<br>(0.3 - 4.3) | 1<br>(0.07 - 10.2) | 0.8<br>(0.06 - 1.9) | 1<br>(0.09 - 1.4) | 0.214 | 0.628 | 0.668 |
| *Phocaeicola* | 1.2<br>(0.3 - 5.2) | 0.9<br>(0.09 - 2.8) | 2.7<br>(0.3 - 4) | 2.2<br>(0.6 - 4.2) | 0.5<br>(0.3 - 0.7) | 0.4<br>(0.4 - 0.5) | 0.036* | 0.430 | 0.619 |
| *Ruminococcus* | 1.1<br>(0.4 - 22.9) | 0.9<br>(0.7 - 19.2) | 0.8<br>(0.1 - 5.3) | 0.8<br>(0.3 - 1) | 1.7<br>(0.2 - 5.3) | 1.2<br>(0.008 - 3.2) | 0.491 | 0.055 | 0.841 |
| *Solibaculum* | 1.1<br>(0.4 - 10.8) | 3.4*<br>(0.8 - 7.8) | 1<br>(0.07 - 6.7) | 1<br>(0.08 - 6.4) | 5.8<br>(0.03 - 8) | 1.4*<br>(0.02 - 3) | 0.782 | 0.172 | 0.046* |
| *Herbinix* | 1<br>(0.3 - 2.8) | 1.6<br>(0.2 - 2.4) | 0.6<br>(0.2 - 1.9) | 0.8<br>(0.1 - 1.5) | 1.1<br>(0.5 - 4.6) | 1.5<br>(0.8 - 3.3) | 0.403 | 0.740 | 0.685 |
| *Lachnoclostridium* | 0.9<br>(0.5 - 2.3) | 0.6<br>(0.4 - 3) | 0.8<br>(0.2 - 1.9) | 0.6<br>(0.3 - 3.5) | 0.6<br>(0.01 - 0.8) | 0.6<br>(0.2 - 2.8) | 0.487 | 0.103 | 0.934 |
| *Anaerostipes* | 0.9<br>(0.3 - 6.7) | 1<br>(0.4 - 7.2) | 0.8<br>(0.3 - 4) | 0.8<br>(0.3 - 1.2) | 0.5<br>(0.2 - 6) | 1<br>(0.01 - 5.7) | 0.801 | 0.617 | 0.806 |
| *Escherichia* | 0.8<br>(0.2 - 2.7) | 0.3<br>(0.1 - 2.9) | 1.3<br>(0.1 - 14) | 1.7<br>(0.2 - 5) | 1.1<br>(0.3 - 19.3) | 3.4<br>(0.3 - 9.2) | 0.012* | 0.291 | 0.756 |

The values are presented as medians and ranges. Based on the median test across time points within groups, \*indicates significant differences (p<0.05).

### 2.3. Species level

Four species dominated the intestinal microbiota of cancer patients: *Faecalibacterium prausnitzii, Anaerobutyricum hallii*, and *Vescimonas coprocola* and *Vescimonas fastidiosa*. For *Bacteroides* sp. PHL 2737 and *Vescimonas coprocola*, we observed significant differences in the interaction between treatment and time; between baseline and 3 months, *Bacteroides* sp. PHL 2737 decreased significantly in both the DMB groups and *Vescimonas coprocola* decreased in the placebo group (Table 3).

**Table 3.**
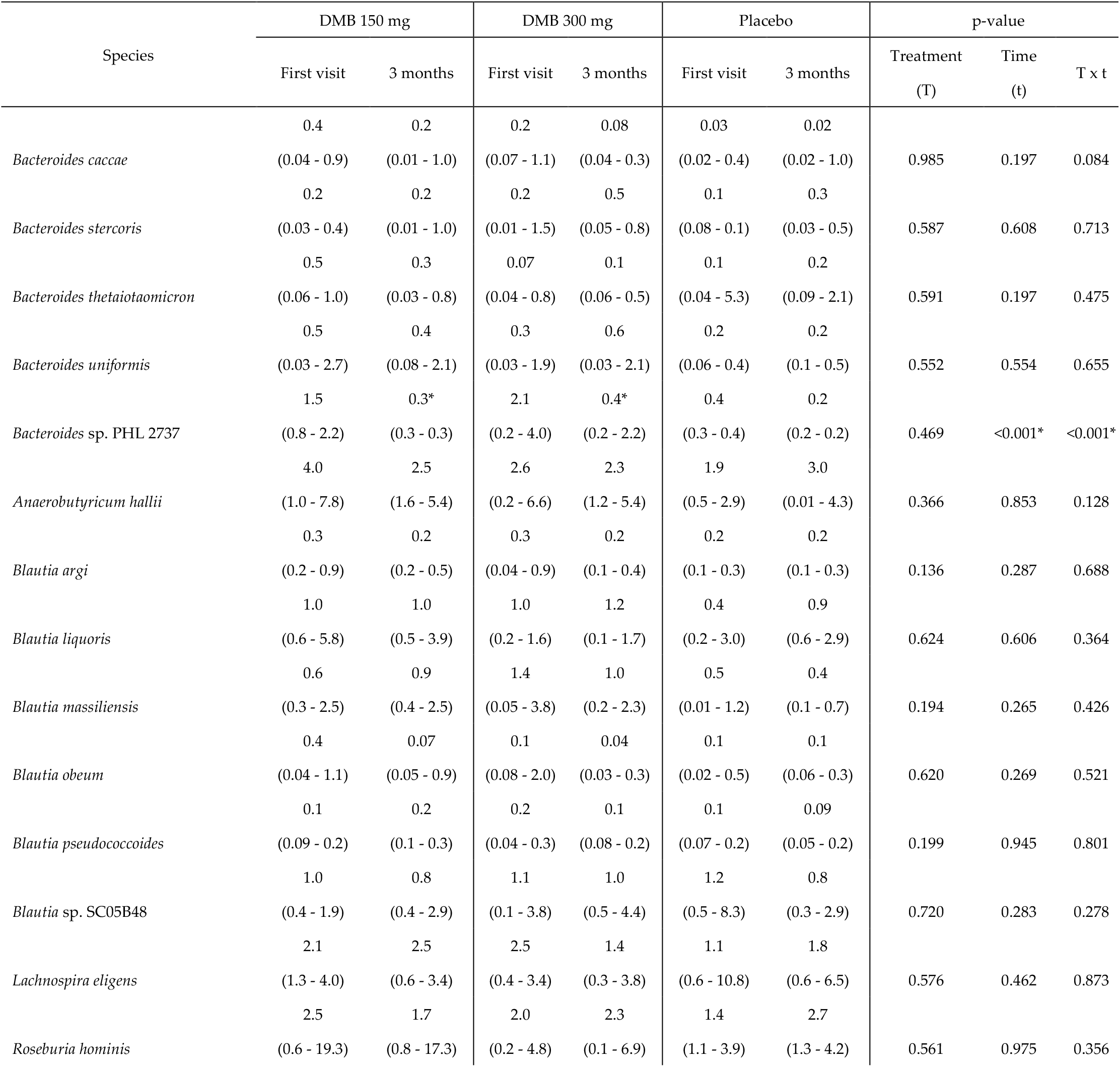
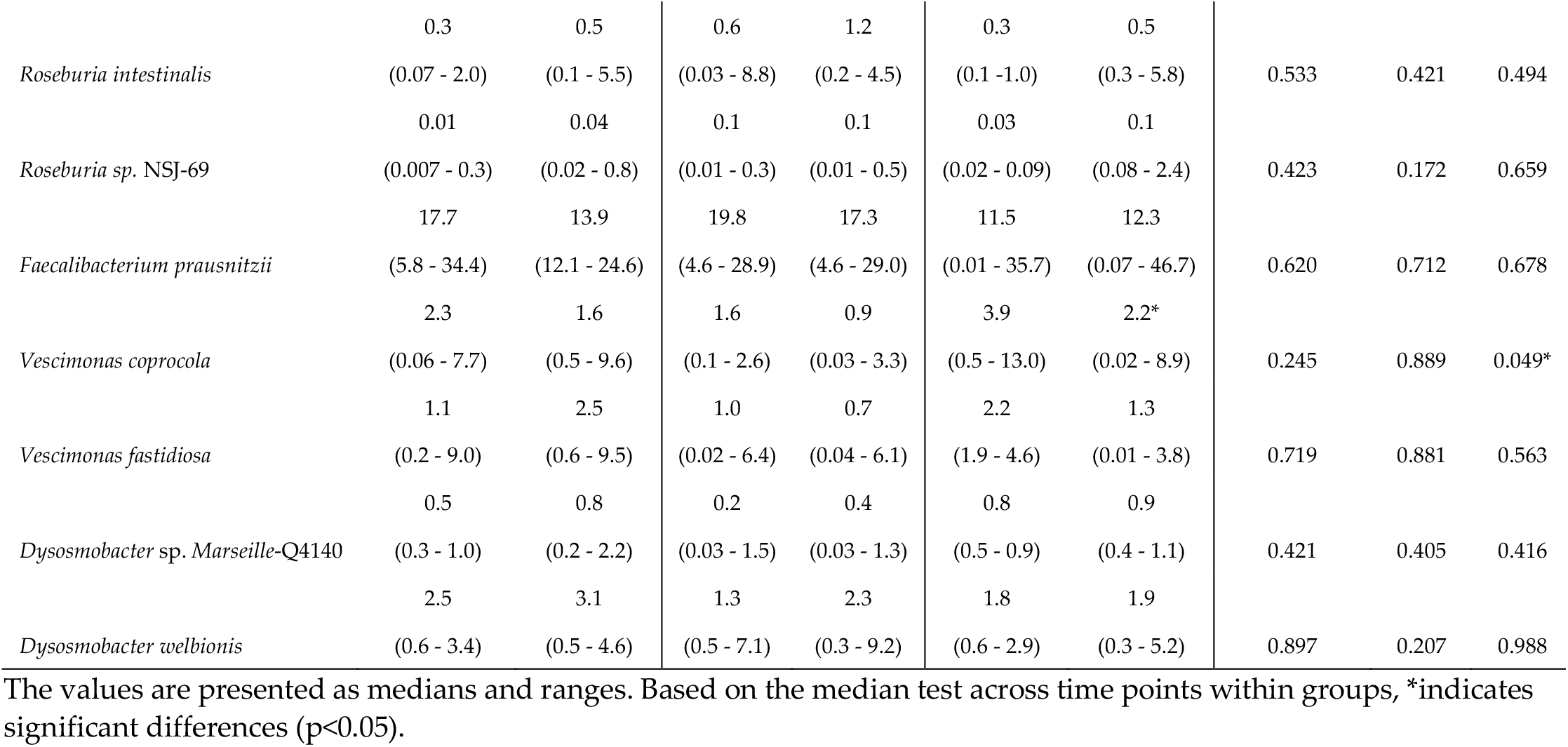
Relative abundances of intestinal bacteria at the species level in malnourished patients with cancer and dysgeusia who received standard-dose DMB (150 mg), high-dose DMB (300 mg) or placebo for 3 months.

### 2.4. Short-chain fatty acids

In all the study groups, acetic acid was the most abundant short-chain fatty acid. For acetic acid there were significant differences between times and the interaction of treatment and time. As a result of treatment with the standard dose of DMB, the acetic acid level increased significantly, whereas the level decreased in patients receiving the placebo treatment (Table 4).

**Table 4.** Plasma short-chain fatty acids in malnourished patients with cancer and dysgeusia who received standard-dose DMB (150 mg), high-dose DMB (300 mg) or placebo for 3 months.

| Short-chain fatty acids | DMB 150 mg |  | DMB 300 mg |  | Placebo |  | p-value |  |  |
| --- | --- | --- | --- | --- | --- | --- | --- | --- | --- |
|  | Baseline | 3 months | Baseline | 3 months | Baseline | 3 months | Treatment (T) | Time (t) | T x t |
| Acetic acid (μmol/L) | 12.8 ± 6.4 | 26.7 ± 6.7* | 36.6 ± 7.0 | 31.7 ± 7.3 | 25.0 ± 7.8 | 15.0 ± 8.2* | 0.082 | 0.032* | 0.027* |
| Propionic acid (μmol/L) | 0.8 ± 0.6 | 2.2 ± 0.9 | 1.5 ± 0.6 | 0.6 ± 1.0 | 1.9 ± 0.7 | 2.6 ± 1.1 | 0.357 | 0.420 | 0.559 |
| Isobutyric acid (μmol/L) | 0.3 ± 0.1 | 0.4 ± 0.1 | 0.3 ± 0.1 | 0.3 ± 0.1 | 0.4 ± 0.1 | 0.4 ± 0.1 | 0.993 | 0.893 | 0.955 |
| Butyric acid (μmol/L) | 0.9 ± 0.3 | 1.3 ± 0.3 | 0.9 ± 0.3 | 0.8 ± 0.4 | 1.1 ± 0.3 | 1.3 ± 0.4 | 0.698 | 0.414 | 0.591 |
| Isovaleric acid (μmol/L) | 0.2 ± 0.1 | 0.3 ± 0.1 | 0.2 ± 0.1 | 0.3 ± 0.1 | 0.2 ± 0.1 | 0.4 ± 0.1 | 0.865 | 0.991 | 0.603 |
| Valeric acid (μmol/L) | 1.0 ± 0.8 | 2.2 ± 0.9 | 1.0 ± 1.5 | 0.7 ± 1.0 | 1.0 ± 1.9 | 2.7 ± 1.1 | 0.573 | 0.559 | 0.878 |

The values are presented as the means and standard deviations. Based on the median test across time points within groups, \*indicates significant differences (p<0.05).

### 2.5. Rivera-Pinto for microbiota balance

To ascertain the microbiome balance at the conclusion of the trial, the Rivera-Pinto method was employed [46]. The analysis revealed that *Pseudomonadota*, was most associated with the placebo group when the standard-dose DMB group (150 mg) was compared with the placebo group (Figure 1A). In the standard-dose DMB group, lower balance scores were associated with lower relative abundances of *Roseburia, Phocaeicola, Escherichia* and *Streptococcus* than *Pseudomonadota* (Figure 1A). With respect to the high-dose DMB group versus the placebo group, *Escherichia* was the most strongly associated with the placebo group (Figure 1B). Thus, the higher the dose of DMB was, the lower the balance scores associated with lower relative abundances of *Actinobacteriota* than of *Escherichia* (Figure 1B).

**Figure 1.**
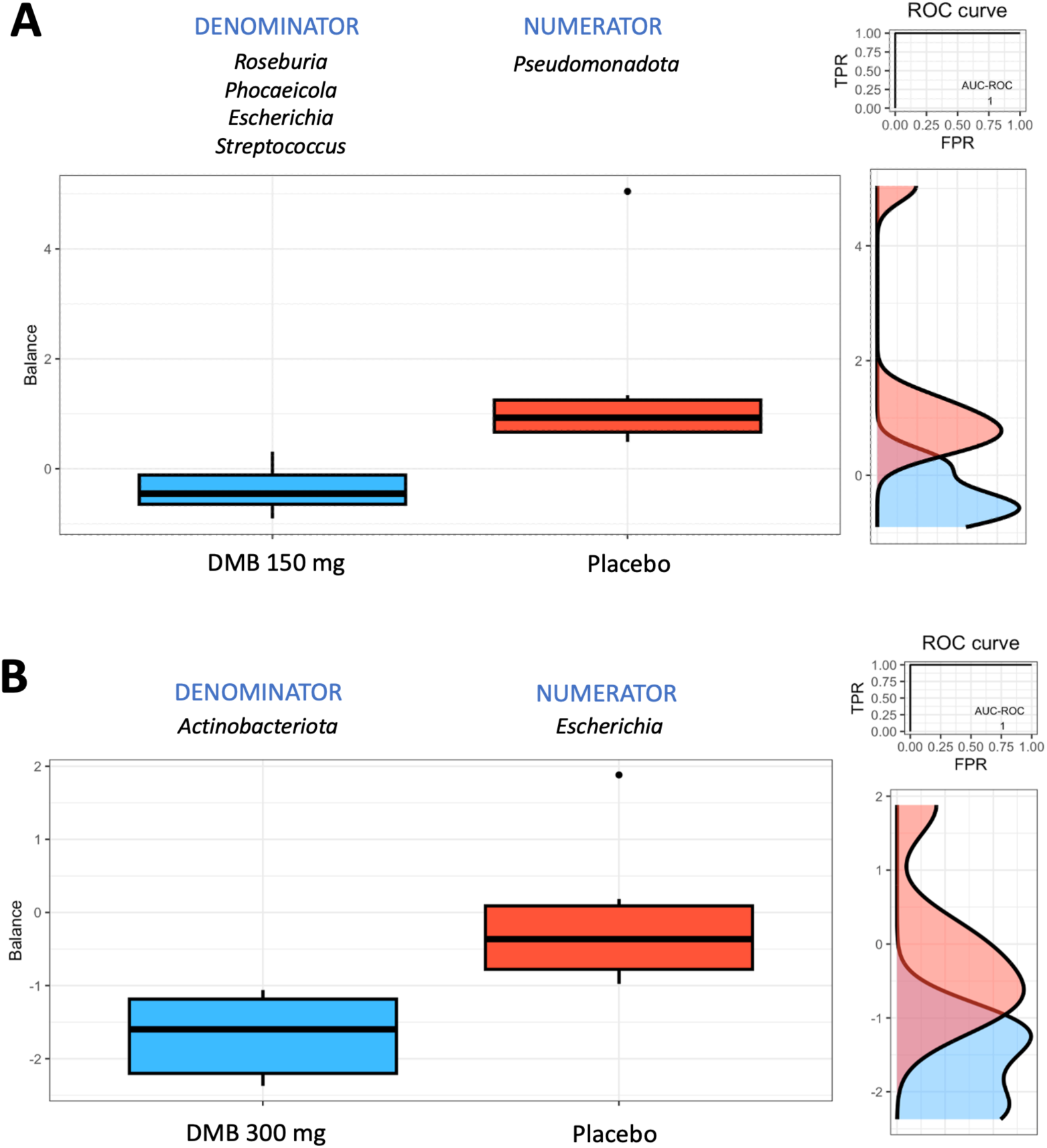
Group balances are presented in an overview. The top of the plot indicates that groups of taxa constitute the global balance. Box plots illustrating the distribution of balance scores for the DMB 150 mg (standard dose) and placebo groups (A) and the DMB 300 mg (high dose) and placebo groups (B). On the right, the ROC curve with its AUC value and the density curve are displayed.

### 2.6. Analysis of the relationships among the intestinal microbiota, nutritional status, electrical taste perception inflammatory cytokines,, an plasma short-chain fatty acids

In the group of patients with cancer and dysgeusia who received the standard dose of DMB, *Mediterraneibacter* had a negative correlation with saturated fatty acid percentage of energy in the diet. TNF-α levels were positively correlated with *Lachnoclostridium* and *Mediterraneibacter*. The presence of the *Prevotella* genus was positively correlated with the electrogustometry values on the right side of the tongue and the proteolysis inducing factor (Figure 2A).

**Figure 2.**
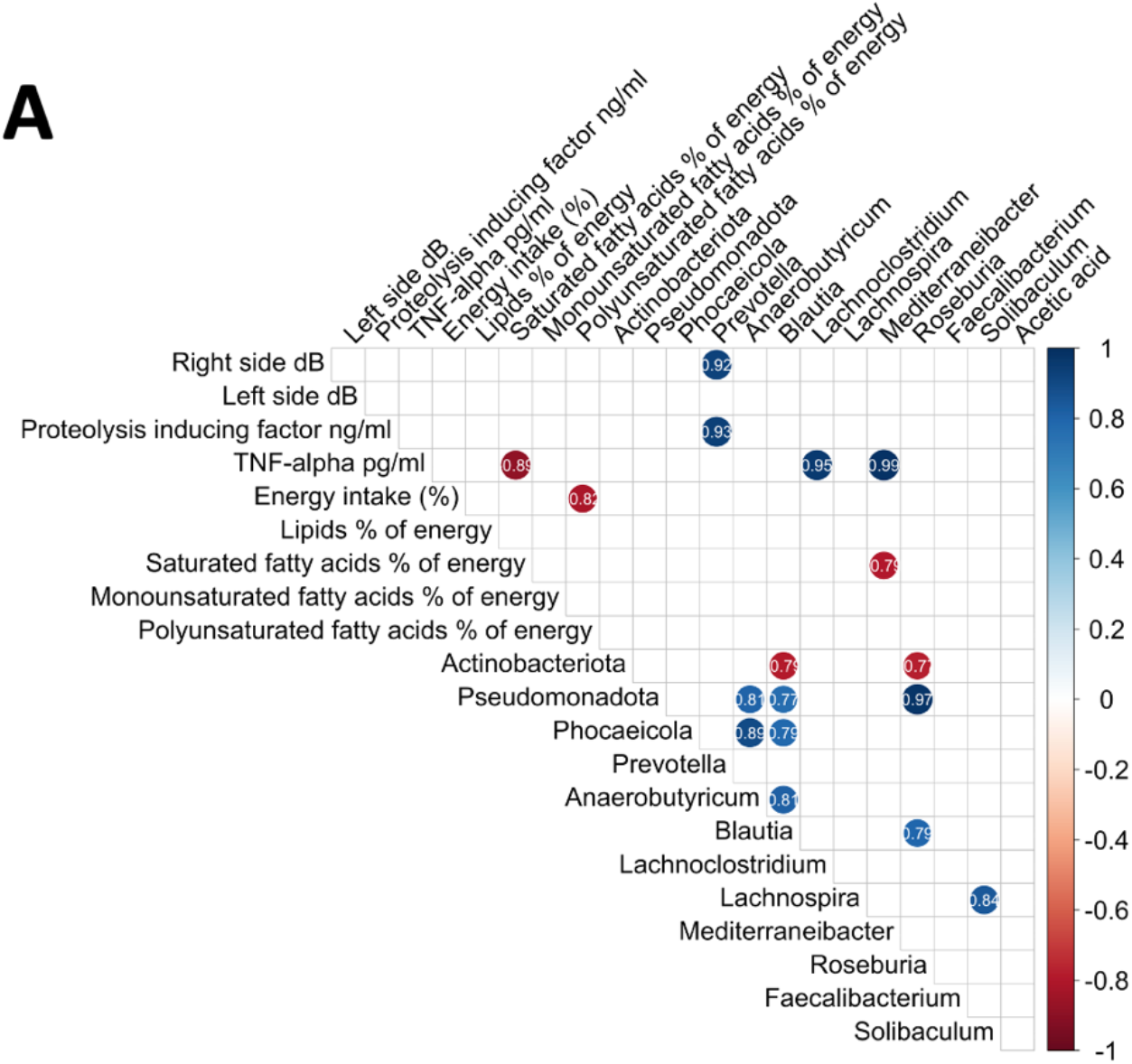
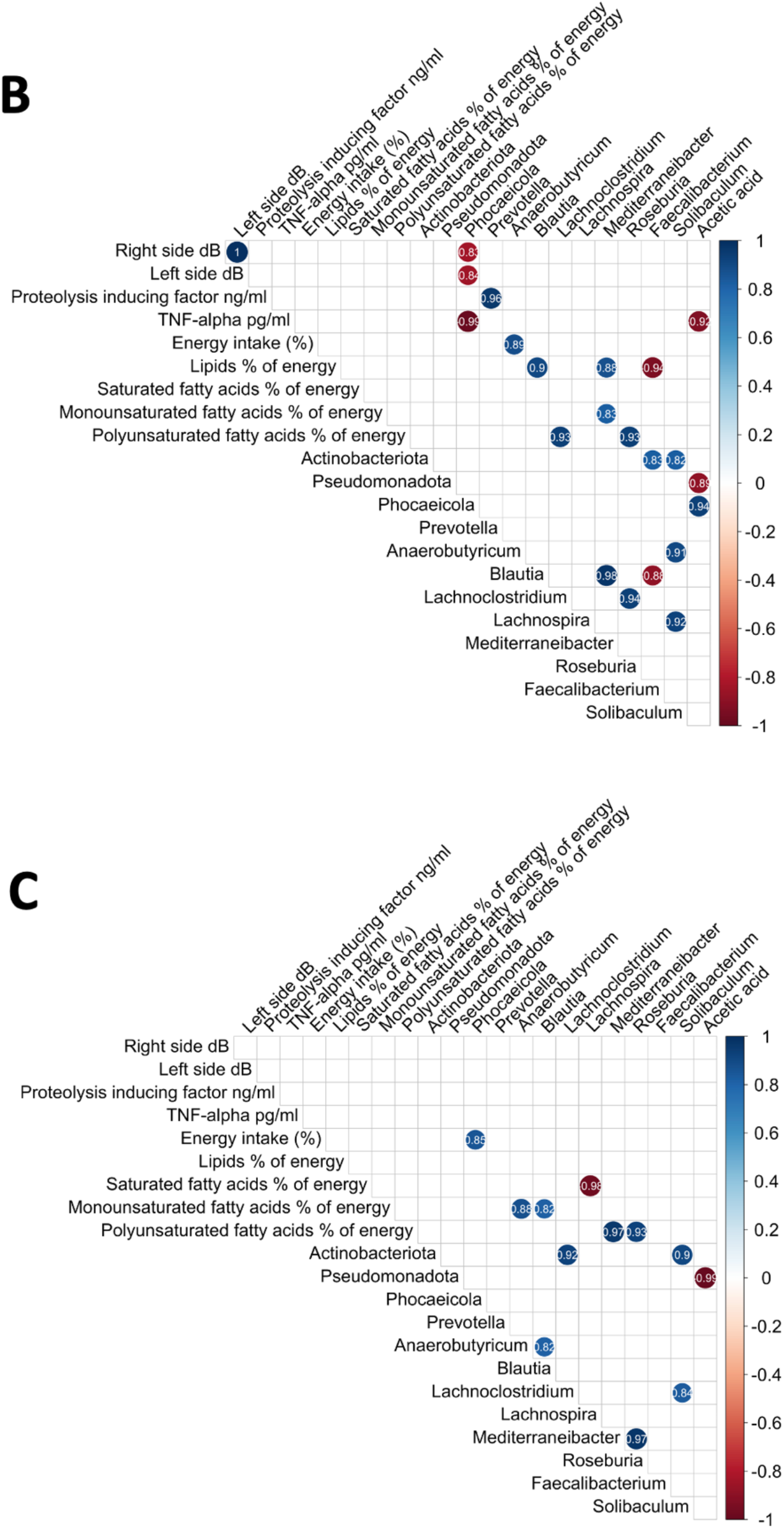
Correlations between the intestinal microbiota, nutritional status, electrical taste perception and inflammatory cytokines. A. DMB 150 mg (standard dose), B. DMB 300 mg (high dose), and C. placebo.

Several correlations were observed in the group that received high doses of DMB (Figure 2B). As a percentage of energy, *Blautia* and *Mediterraneibacter* were positively associated with lipids in the diet, whereas *Faecalibacterium* was negatively associated. *Mediterraneibacter* was positively correlated with dietary monounsaturated fatty acids. There was a positive correlation between dietary polyunsaturated fatty acids (PUFAs) and *Lachnoclostridium* and *Roseburia*. Dietary energy intake (%) was positively correlated with *Anaerobutyricum*. There was a negative correlation between the relative abundance of *Phocaeicola* and electrogustometry (both, right and left sides of the tongue), and TNF-α levels. Proteolysis inducing factor was positively correlated with the *Prevotella* genus. There was a positive correlation between *Phocaeicola* and plasma acetic acid concentration, whereas a negative correlation was detected between *Pseudomonadota* and TNF-α levels (Figure 2B).

A positive association between *Phocaeicola* and energy intake was detected in the placebo group. Dietary saturated fatty acids (%) were negatively associated with *Lachnospira*. Dietary monounsaturated fatty acids were positively correlated with *Anaerobutyricum* and *Blautia*. Dietary PUFAs were positively correlated with *Mediterraneibacter* and *Roseburia* (Figure 2C).

## 3. Discussion

The present study revealed that regular DMB consumption together with nutritional treatment and individualized dietary advice changed the composition of the gut microbiota. We found that the differences between the genera *Phocaeicola* and *Escherichia* were dependent on the treatment, but only the *Solibaculum* genus presented an increased relative abundance in the standard-dose DMB group following 3 months. After 3 months, *Bacteroides* sp. PHL 2737 showed greater relative abundance in both DMB groups, whereas *Vescimonas coprocola* was more abundant in both treatment groups. According to the electrogustometry results on the right side of the tongue of patients with cancer and dysgeusia receiving the standard dose of DMB, the presence of *Prevotella* genus was positively correlated with the electrogustometry values and proteolysis-inducing factor plasma levels. The TNF-α levels were positively correlated with *Lachnoclostridium* and *Mediterraneibacter*. The abundance of *Mediterraneibacter* was negatively correlated with dietary saturated fatty acids expressed as percentage of the dietary energy. In the group that received high doses of DMB, several correlations were observed. A negative correlation was found between the relative abundance of *Phocaeicola* and electrogustometry (both right and left sides of the tongue) as well as TNF-α levels. The proteolysis inducing factor was positively correlated with the *Prevotella* genus. *Anaerobutyricum* was positively correlated with the energy intake. *Blautia* and *Mediterraneibacter* were positively associated with lipids in the diet, whereas *Faecalibacterium* was negatively associated. The correlation between *Mediterraneibacter* and monounsaturated fatty acids was positive. *Lachnoclostridium* and *Roseburia* were positively correlated with dietary PUFAs. Also, our results revealed that *Phocaeicola* was positively correlated with the plasma acetic acid concentration, whereas *Pseudomonadota* was negatively correlated with the TNF-α levels.

According to Hes et al. (2024), the gut microbiome of patients with severe mucositis differ from that of patients with grades 1-2 mucositis, with an increase in the abundances of *Mediterraneibacter (Ruminococcus gnavus*) and *Clostridiaceae*, including *Hungatella hathewayi* [47]. As shown here, the habitual consumption of a standard dose of DMB was positively associated with TNF-α levels and *Lachnoclostridium*, and *Mediterraneibacter* abundances, whereas a high dose of DMB was negatively associated with TNF-α levels and the relative abundance of *Phocaeicola*. Microbes, as well as gut bacteria-derived metabolites, pathogen-associated molecular patterns, and antigens, can move from the gastrointestinal tract to other closely related tissues and impact cancer progression [48].

The *Bacteroides* and *Phocaeicola* species play crucial roles in the human colon. By degrading complex heteropolysaccharides into short-chain fatty acids, those organisms contribute to the body’s use of these compounds [49]. Our findings indicate that the consumption of DMB at high doses is positively correlated with the abundance of the genus *Phocaeicola* and acetic acid concentrations. In addition, following DMB administration at a high dose, a positive correlation was found between PUFAs, *Lachnoclostridium*, and *Roseburia*. PUFAs have antitumor activity. In particular, it has been proposed that PUFAs, specifically eicosapentaenoic acid and docosahexaenoic acid, possess anticolorectal cancer activity [50]. A recent study investigated the impact of PUFA supplementation on the fecal microbiome in middle-aged, healthy volunteers, showing that PUFA supplementation leads to a reversible increase in bacteria that produce short-chain fatty acids [51]. Moreover, a reversible increase in the abundance of several bacterial genera, including *Bifidobacterium, Roseburia* and *Lactobacillus*, was observed in patients who received one or both PUFA treatments. Consequently, a high dose of DMB may enhance the presence of microorganisms that increase SCFA availability and contribute to PUFA consumption.

Numerous diseases in humans have been associated with changes in the gut microbiota composition, with fluctuations in the prevalence of particular bacterial groups. In this regard, *Faecalibacterium* is one of the most notable genera. The relative abundance of this bacteria was estimated to be between 11 and 20%. A recent study revealed a negative correlation between the abundance of *Faecalibacterium* and increased intraindividual variability in microbiota composition, indicating that it as a keystone taxon [52,53].

Certain species of *Faecalibacterium* have been observed to undergo alterations in a number of diseases and disorders. In fact, multiple studies have demonstrated that a high baseline level of *Faecalibacterium*, along with that of other *Bacillota*, is positively correlated with responses to related treatments for various cancers, including melanoma [54-58], hepatocellular carcinoma [59] and non-small cell lung cancer [60]. We observed a tendency to decrease concentration of *Faecalibacterium prausnitzii* in DMB groups.

We showed that standard dose of DMB administration resulted in a significant increase in the relative abundance of *Solibaculum* genus, whereas placebo resulted in a reduction of this genus. We identified four species of microbes that are dominant within the gastrointestinal tract of patients with cancer: *Faecalibacterium prausnitzii, Anaerobutyricum hallii, Vescimonas* species, *V. coprocola* and *V. fastidiosa*. However, we identified significant differences in the interaction effect between treatment and time for *Bacteroides* sp. PHL 2737 and *Vescimonas coprocola*. A significant decrease in *Bacteroides* sp. PHL 2737 levels was observed between the baseline and the three-month period in both the DMB groups. Additionally, *Vescimonas coprocola* levels decreased significantly in the placebo group. Environmental alterations caused by dysbiosis can result in a gradual decline of functional redundancy, either as a consequence of disease or its treatment. A recent study demonstrated that colorectal cancer is influenced by the co-occurrence of species, including *Vescimonas coprocola* and *Vescimonas fastidiosa*, although they did not significantly differ in abundance [61]. Metagenomic studies based on colorectal cancer datasets have reported an association between specific microbial species and this type of cancer [62-70]. For example, multiple studies have demonstrated the main role of particular species in the development of colorectal cancer [71], such as *Streptococcus gallolyticus* [72], *Bacteroides fragilis* [73,74] and *Fusobacterium nucleatum* [75-77]. Moreover, it has been proposed that *Bacteroides fragilis* [73,74] and *Fusobacterium nucleatum* [75-77] are key factors in the tumorigenesis process and then they may be replaced by “passenger” species that are favored by the cancer microenvironment [78].

The intake of DMB together with nutritional treatment and individualized dietary advice results in positive changes in the intestinal microbiome of cancer patients and patients with dysgeusia, which are correlated with taste perception in the DMB high dose group. By utilizing these strategies, patients with cancer are able to maintain their nutritional intake and enjoy their meals despite changes in taste perception and aftertaste [79]. It is, however, important for patients to discuss their dietary preferences and modifications with healthcare professionals in order to ensure that they receive adequate nutrition while undergoing cancer treatment [79]. The ability to manipulate gut microbiome composition to improve cancer therapy outcomes is a significant new area of research [80,81]. Intestinal microbiome composition is susceptible to changes due to diet and the environment, so educating patients on food consumption during cancer treatment and avoiding carcinogens may improve outcomes [80,81].

Cancer patients receiving a standard dose or a higher dose of DMB experienced different changes in their gut microbiota from those receiving a placebo. The difference could be attributed to the sweet taste experienced following the ingestion of orodispersible DMB tablets before each main meal, compared to the placebo group, which may lead to a better dietary intake.

## 4. Materials and Methods

### 4.1. Statement of ethical principles

This project was approved by University Hospital La Paz’s (HULP Code 6164) Scientific Research and Ethics Committee in June 2022. According to the Declaration of Helsinki’s Ethical Standards, this study adheres to recommendations for physicians conducting biomedical research on humans. The ICH Harmonized Tripartite Guidelines should be familiarized and followed by all researchers in order to maintain good clinical practices.

The research team informed the patients (verbally and in writing) of the study characteristics and the responsibilities of participation in the trial before they signed the informed consent form. Patients were informed during the study that they could withdraw from the study at any time by notifying their doctor without giving a reason. The processing of personal information is subject to several legal requirements, including Spanish Organic Law 3/2018 of 5 December and the General Data Protection Regulation of the European Union (EU) 2016/679 of 27 April 2016.

### 4.2. Participants and experimental design

Detailed information of CLINMIR study is published elsewhere [43,45]. Briefly, the CLINMIR study is a pilot randomized, parallel, triple-blind, and placebo-controlled clinical trial. Using the number NCT05486260, the present protocol was registered at http://clinicaltrials.gov, accessed on 14 March 2024. An oncology service and clinical nutrition unit at HULP in Madrid recruited 31 malnourished cancer patients with taste disorders.

Three treatment arms were randomly assigned to malnourished patients with cancer and taste disorders who were receiving active treatment. A miraculin-based food supplement was administered to patients five minutes prior to each meal (breakfast, lunch, and dinner) during a three-month study. The tablets contained either DMB at one of its two dosages or a placebo [43,45].

Each intervention group consisted of ten patients who were randomly assigned to receive one of two DMB dosages or a placebo. In the first arm of the study, 150 mg of DMB equivalent to 2.8 mg of miraculin is combined with 150 mg of freeze-dried strawberries; in the second arm, 300 mg of DMB is utilized equivalent to 5.5 mg of miraculin; and in the third arm, 300 mg of freeze-dried strawberries are used as a placebo. Each of the three treatments was isocaloric (Table S1). The subjects received as many tablets as necessary during scheduled visits to the HULP in order to complete the three-month intervention period [43,45].

### 4.3. Sequencing of biological samples

To prepare for the analyses, sterile plastic containers were used to collect fecal samples at baseline and 3 months after intervention. Blood samples were collected by trained personnel at the HULP Extraction Unit in the morning (approximately at 8:00 am) during blood tests before chemotherapy to avoid unnecessary punctures and hospitalizations. The blood samples were collected in vacuum tubes, labeled, transported, and centrifuged at 1500 x g for 10 minutes. We prepared and labeled aliquots of blood samples according to a numerical code and stored them at −80 °C.

#### 4.3.1. Extraction of DNA

QIAamp Fast DNA Stool Mini kit (ref. ID: 51604, Qiagen Inc., Hilden, Germany) was used to extract DNA from the stool samples. The purity and integrity of DNA were determined using spectrophotometry (NanoDrop, Thermo Fisher Scientific, Massachusetts, USA).

#### 4.3.2. 16S rRNA gene sequencing and taxonomic assignment

A detailed description of 16S sequencing via Oxford Nanopore Technologies can be found elsewhere [44]. Briefly, the 16S rRNA gene was PCR-amplified using redesigned 16S primers (27F and 1492R) with 5’ tags that facilitate ligase-free attachment. By vortexing 30 μl of AMPure XP beads (Beckman Coulter, ThermoScientific, Spain), and mixing by pipetting, PCR products from each sample were cleaned.

In order to achieve a concentration of 50-100 fmoles, all barcoded libraries were combined in the appropriate ratios. The final library was loaded into the SpotON Flow Cell Mk R9 Version (ref. FLO-MIN106D, Oxford Nanopore Technologies, Oxford, United Kingdom) using the Minion M1kc and M1kb sequencers (Oxford Nanopore Technologies, Oxford, United Kingdom).

After the raw data had been generated, searches were performed via Guppy version 6.5.7 (Oxford Nanopore Technologies, Oxford, United Kingdom), and sequences were identified using Kraken2 (refseq Archaea, bacteria, viral, plasmid, human, UniVec_Core, protozoa, fungi & plant database) and further analyzed using QIIME2 [82]. Assigning taxonomy to ASVs was performed using the classify sklearn naïve Bayes taxonomy classifier (via q2-feature-classifier) [83] using SILVA 16S V3-V4 v132_99 [84] with a similarity threshold of 99%. The diversity of the samples was examined using the vegan library [85]. In this study, Shannon, Simpson and Chao1 indices were examined.

### 4.4 Plasma cytokines

Plasma tumor necrosis factor-alpha (TNF-α), and human proteolysis-inducing factor (PIF) were analyzed as previously described [86].

### 4.5 Dietary pattern assessment

For three days, including one holiday, daily food records were kept. Patients were advised to record household measurements (spoonfuls, cups, etc.) or household weights in the absence of weight records. A nutritionist reviewed all records in the presence of the patient to ensure that the information collected was accurate and complete. DIAL software (Alce Ingeniera, Madrid, Spain) was used to convert the energy and nutrients contained in foods, drinks, dietary supplements, and preparations. Finally, the results were compared with the recommended intakes for the Spanish population [43,45].

### 4.6 Short-chain fatty acids determination by gas chromatography/mass spectrometry

One hundred μl of plasma were individually placed in 1.5 ml tubes. Afterward, 10 μL of acidified water (15 % phosphoric acid v/v), and 10 μl of internal standards (sodium acetate ^13^C2 at 300 μM, butyric-1,2-^13^C2 at 60 μM and isobutyric acid d6, valeric acid d9, isovaleric acid d9 and propionic d6 acid at 30 μM) are added and vigorously mixed up. Next, a liquid-liquid extraction was performed with 150 μl of MTBE. The extraction was assisted by vortexing for 10 minutes. At this point, tubes were centrifuged at 15000 rpm for 10 minutes at 4 ºC. One hundred μl were transferred into a vial with an insert. The vials were centrifuged at 1000 rpm for 30s at 4 ºC and 1 μl was injected into gas chromatography/mass spectrometry. Briefly, short-chain fatty acids were separated on a DBFFAP chromatographic column (30 m x 0.25 mm x 0.25 μm). The oven temperature was programmed as follows: (i) initial temperature 40 ºC, (ii) linearly increased at 12 ºC/min until 130 ºC (0 min), (iii) then linearly raised at 30 ºC/min to 200 ºC (0 min) and (iv) in the final step the temperature was ramped at 100 ºC/min to 250 ºC (4.5 min). The column flow was set at 1.5 ml/min with Helium as carrier gas. The injector was set at 250 ºC and the extract was injected in a split-less mode. Using electronic impact (70 eV) for ionization, the mass analyzer was operated for multi reaction monitoring.

### 4.7 Statistical analysis

To examine the effects of time, treatment, and their interaction (time x treatment), a linear mixed model was used to examine the differences between placebo, 150 mg of DMB, and 300 mg DMB. Using the R program, a linear mixed model was developed using the lme4 package [87]. A median test revealed significant differences across time points within groups.

We also examined the relationships between intestinal microbiome variables, inflammatory parameters, dietary variables, short-chain fatty acids, and electrical taste perception outcomes via Pearson’s correlations; for that purpose we used the R Studio’s corrplot function [88] correcting multiple testing using the FDR procedure [89]. Only significant and corrected associations are shown in the graphs. The red and blue lines in the graphs indicate correlation values, with negative correlations highlighted in red (−1) and positive correlations highlighted in blue (+1).

Rivera-Pinto analysis can identify microbial signatures, i.e., groups of microbes capable of predicting particular phenotypes of interest. This microbial signature may be used to diagnose, prognosticate, or predict therapeutic response on the basis of the unique microbiota of an individual. Identifying microbial signatures requires modeling the response variable and selecting the taxa that are the most accurate at classification or prediction. To select a sparse model that adequately explains the response variable, we evaluated specific signatures at the phylum and genus levels using the Rivera-Pinto method and the Selbal algorithm. Based on data collected from two groups of taxa, microbial signatures were calculated using geometric means. These groups are those with relative abundances or balances that are related to the response variable of interest [46].

## 5. Conclusions

This pilot randomized, parallel, triple-blind, and placebo-controlled clinical trial identified a putative innovative therapeutic option for the management of taste disorders in patients with cancer. This novel strategy was designed with the intent of reducing the adverse effects associated with chemotherapeutic, radiotherapeutic, and immunotherapeutic interventions, which may include alterations in taste, changes in body composition and nutritional status, and alterations in the quality of life [45]. Here, we observed differences between the genera *Phocaeicola* and *Escherichia* depending on the treatment. Only the *Solibaculum* genus increased in relative abundance in the DMB group after 3 months. With respect to species, *Bacteroides* sp. PHL 2737 had a lower relative abundance in both DMB groups, and *Vescimonas coprocola* exhibited a greater abundance in both treatments after 3 months. Moreover, a standard dose of DMB was positively associated with TNF-α levels and *Lachnoclostridium* and *Mediterraneibacter* abundances, whereas a high dose of DMB was negatively associated with TNF-α levels and the relative abundance of *Phocaeicola*. After high-dose DMB administration, a positive correlation was found between PUFAs, *Lachnoclostridium*, and *Roseburia*. Additionally *Phocaeicola* was positively correlated with acetic acid levels. Accordingly, DMB intake and nutritional treatment positively modify the intestinal microbiome in patients with cancer and dysgeusia, which might lead to a greater immunological response and better dietary intake.

## Supplementary Materials

The following supporting information can be downloaded at: www.mdpi.com/xxx/s1, Table S1: Nutritional composition of the food supplement enriched in miraculin (DMB) and placebo.

## Author Contributions

Conceptualization, B.L.-P., A.G. and S.P.-M.; methodology, B.L.-P. and J.F.-B.; software, J.D.-P.; validation, F.J.R.-O, A.I.A.-M. and M.B.-H.; formal data analysis, J.D.-P., F.J.R.-O, and M.B.-H.; investigation, B.L.-P and L.A.-C.; resources, S.P.-M.; data curation, L.A.-C; writing-original draft preparation, J.D.-P., F.J.R.-O, M.B.-H. and A.G.; writing-review and editing, J.D.-P., F.J.R.-O, M.B.-H. and A.G.; supervision, S.P.-M and A.G.; project administration, B.L.-P; funding acquisition, S.P.-M. All authors have read and agreed to the published version of the manuscript.

## Institutional Review Board Statement

The study was conducted under the Declaration of Helsinki, and approved by the Ethics Committee of La Paz University Hospital (protocol code 6164, 23 June 2022).

## Informed Consent Statement

Informed consent was obtained from all subjects involved in the study.

## Data Availability Statement

A reasonable request should be made to the corresponding author for access to the datasets used and/or analyzed in the current study.

## Acknowledgments

J.P.-D. is part of the “UGR Plan Propio de Investigación 2016” and the “Excellence actions: Unit of Excellence in Exercise and Health (UCEES), University of Granada”. F.J.R.-O. is supported by a grant from the Spanish Government’s “Agencia Estatal de Investigación-Juan de la Cierva-Incorporación” program (IJC2020-042739-I). We thank (Lucía Tadeo, Helena Torrell, Adría Cereto and Núria Canela) from the Genomics facility of the Centre for Omic Sciences (COS) Joint Unit of the Universitat Rovira i Virgili-Eurecat, for their contributions to the sequencing analysis.

