## Supplementary material for "Effect of a novel food rich in miraculin on the intestinal microbiome of malnourished patients with cancer and dysgeusia": Table S1

**Table S1**. Nutritional composition of the food supplement enriched in miraculin (DMB) and placebo

| | | Standard dose of DMB (150 mg DMB + 150 mg strawberry freeze-dried) | High dose of DMB (300 mg DMB) | Placebo (300 mg strawberry freeze-dried) |
|---|---|---|---|---|
| Energy | kcal | 0.99 | 1 | 0.97 |
| Carbohydrates | mg | 194 | 234 | 154 |
| Sugars | mg | 156 | 162 | 150 |
| Fiber | mg | 26 | 6 | 46 |
| Proteins | mg | 20 | 15 | 24 |
| Lipids | mg | 9 | 5 | 12 |
| Saturated fatty acids | mg | 2 | 2 | 1 |
| Sodium chloride | mg | 0.1 | 0.1 | 0.03 |
| Humidity | mg | 4 | 4 | 5 |
| Ash | mg | 12 | 14 | 15 |
| Miraculin | mg | 2,8 | 5,6 | 0 |

Nutritional composition provided by Medicinal Gardens, S.L.
